# The GenoDiabMar registry. A collaborative research platform of “real world” type 2 diabetes patients

**DOI:** 10.1101/2021.10.12.21264882

**Authors:** Adriana Sierra, Sol Otero, Eva Rodríguez, Anna Faura, María Vera, Marta Riera, Vanesa Palau, Xavier Durán, Anna Costa-Garrido, Laia Sans, Eva Márquez, Vladimir Poposki, Josep Franch-Nadal, Xavier Mundet, Anna Oliveras, Marta Crespo, Julio Pascual, Clara Barrios

## Abstract

The GenoDiabMar registry is a prospective study aims to provide data on demographic, biochemical and clinical changes, from a “real-world” population of Type 2 DM (T2D) patients. This registry is addressed to find new biomarkers related to the micro and macrovascular complications of T2D, especially focused on diabetic nephropathy. The registry includes longitudinal serum and urine samples, DNA bank, as well as data on 227 metabolomics profiles, 77 Immunoglobulin G glycomics traits and others emerging biomarkers. 650 patients aged 69.56 ±9.31 with different grades of chronic kidney disease; (G1-2 50.3%, G3 31.4%, G4 10.8% and G5 7.5%) were followed up for 4.96 (±0.43) years. Regardless of albuminuria, women lost 0.93 (0.40-1.46) glomerular filtration units per year less than men. 17% of the participant experienced rapid progression of renal function, 75.2% men, with differential risk factors between sexes; severe macroalbuminuria >300mg/g for men OR[IQ] 2.40 [1.29:4.44] and concomitant peripheral vascular disease 3.32 [1.10:9.57] for women. An overall mortality of 23% was detected (38% due to Cardiovascular aetiology).

This cohort is postulated as a great tool for scientific collaboration for studies, whether they are focused on T2D, or whether they are interested in comparing differential markers between diabetic and non-diabetic populations.

## 1. INTRODUCTION

Diabetes mellitus affects more than 450 million people in the world, reaching pandemic levels, being the type 2 diabetes (T2D) patients up to 95% of the cases (1). The main complications of diabetes, both microvascular (retinopathy, neuropathy, nephropathy) and macrovascular (ischemic cardiopathy, peripheral vascular disease, cerebrovascular disease), increase the costs associated and entail an important reduction in the quantity and the quality of patients’ life (2–4).

In recent decades, the improvement in prevention strategies and therapeutic interventions has led to a significant reduction in most diabetes complications. However, this is not so evident in the case of diabetic kidney disease (DKD), that remains the leading cause of end stage renal disease (ESRD) in Western countries (5,6). When diabetes induces renal damage, patients have a higher risk of suffering from endothelial disease in any other territory of the body, and patients with DKD have the highest cardiovascular (CV) risk and mortality among patients with chronic kidney disease (CKD) (7–12). Therefore, it is essential to focus attention and effort on the early prevention, diagnosis, and treatment of DKD. Despite the high prevalence and increasing incidence of this disease, the underlying pathophysiological mechanisms are not fully understood and even currently, highly sensitive and specific diagnostic tests are not available. In this way, classical biomarkers used to estimate glomerular filtration rate (eGFR) and renal damage such as serum creatinine and albuminuria, have well-known limitations (13–18), and may fail in the early detection of kidney impairment.

In past years, high-throughput techniques have shown the feasibility of finding new biomarkers of early kidney dysfunction, as well as providing valuable information on the metabolic pathways involved in the physiopathology of DKD and other diabetic micro and macrovascular complications (19–28).

The analysis of hundreds of metabolites, protein glycosylation profiles, genetic variants or emerging biomarkers, require of large sample size cohorts to robustly detect associations. GenoDiabMar is a detailed cohort useful as a tool to improve and expand knowledge on different pathophysiological pathways involved in diabetic complications and allow to replicate results obtained in different populations to generate collaborative research

We present the GenoDiabMar registry, created with the aim of providing data on demographic, biochemical and longitudinal clinical changes, as well as, to obtain biological samples for biobank, from a population of T2D patients attending a real medical out-patient consult. Also, this registry is addressed to find new emerging biomarkers related to the micro and macrovascular complications of T2D, especially focused on diabetic nephropathy.

## 2. METHODS

### 2.1. Study design

The GenoDiabMar registry was designed as a prospective study and currently collected information regarding 650 Caucasians adults with T2D recruited from the nephrologist consultant of Hospital del Mar and six primary care centres from the Hospital del Mar health area, Litoral-Mar of Barcelona, Spain. The inclusion criteria were adults over 45 years old, diagnosed with T2D at least 10 years before the first study visit, if there was no renal disease, and at any time, if renal damage was present. Information on the diagnosis of T2D was retrieved from the patient’s electronical medical records. To avoid the inclusion of patients with pre-diabetes or impaired glucose tolerance and to ensure the T2D diagnosis, only patients under antidiabetic drugs were registered.

Patients with autoimmune polycystic kidney disease or with previously known autoimmune diseases as vasculitis or systemic lupus erythematosus were excluded. Thus, patients who agreed to participate, met the inclusion criteria, and signed the informed consent were enrolled.

Between February 2012 and July 2015, 650 T2D patients underwent a basal in-person medical visit (V1) performed by a nephrologist and a nurse. Medical history, demographics, physical examination, and laboratory data were registered along with collection of blood and urine samples. In addition, an annually follow up of all participants included at the baseline visit was performed to obtain complete analytical and clinical parameters, including new cardiovascular events, changes in the status of diabetic retinopathy and nephropathy, and in mortality, by consulting participants’ electronic clinical reports.

Between March 2017 and February 2020, living patients with functioning kidneys who did not require renal replacement therapy, underwent the second in-person visit (V2). Again, analytical, and clinical data, including changes in treatments, were registered. The second biological samples for biobank were collected in this second visit, performed on an average of 4.96 (±0.43) years from the baseline visit.

The study protocol was approved by the local Medical Ethics Committee of our research institute and the Steering Committee of the Primary Care Area. Patients were previously informed of the collection of samples for the biobank and the use of their clinical data. National guidelines (Code of ethics of the professional association) and international guidelines (Declaration of Helsinki Fortaleza, Brazil, October 2013) were followed. Also, confidentiality of the data was guaranteed in accordance with current regulations: Organic Law 3/2018, of December 5, Protection of Personal Data and guarantee of digital rights and Regulation (EU) No 2016/679 of the Parliament and Council of April 27, 2016 on Data Protection (RGPD). In addition, the study was carried out in accordance with the Biomedical Research Law (Law 14/2007). No binding data are shared for the patient and all data are pseudo-anonymized with a new coding of the medical record number and anonymous labelling of blood samples.

### 2.2. Data Registry

#### Medical records and CV risk factors assessment

Each participant completed a comprehensive questionnaire about their medical history, including information related to the presence and type of Diabetes Mellitus in the family history. Smoking status was registered as current smoker, previous smoker (smoke-free for more than 1 year) or no smoker. Body mass index (BMI) was calculated as weight/height^2^ (Kg/m^2^). The history of CV events was recorded at the baseline and annually and comprised, 1-ischemic heart disease (acute myocardial infarction, angina, cardiac revascularization); 2-cerebrovascular disease (cerebrovascular accident or transient ischemic attack) and 3-peripheral vascular disease (intermittent claudication, ischemic vascular ulcers, or surgical revascularization).

Hypertension was considered if the patient had previously been diagnosed or if they were under anti-hypertensive treatment. In addition, new cases of hypertension were identified if the patient presented systolic blood pressure (SBP) of 140 mmHg or higher, and/or diastolic blood pressure (DBP) of 90 mmHg or higher at the visit (29). Blood pressure was measured with an automatic calibrated sphygmomanometer following a standardized protocol, recording the average of three measurements separated each by 3-5 min. Also, absolutes values were recorded in the dataset.

Dyslipidaemia was considered both as quantitative variable including the absolute values of the lipid profiles component, i.e., determination of total cholesterol> 250mg/dL, LDL cholesterol> 130 mg/dL, HDL cholesterol <35/45 mg/dL (men/women respectively), triglycerides>200 mg/dL (30), and as qualitative variable if previously diagnosed by a physician or the patient used of lipid-lowering medication.

The presence or absence of diabetic retinopathy (DR) is recorded as categorical variable and was diagnosed by fundoscopy performed by an ophthalmologist. It was classified as unknown retinopathy in the case of lack of fundoscopy. In addition, the presence of cataracts diagnosed in the ophthalmological examination was recorded.

The registry also gathered the medication in use at baseline and changes during the follow up, including: Anti-hypertensive drugs distinguishing inhibitors of renin-angiotensin-aldosterone system, calcium antagonists, beta-blockers, diuretics, or combinations; Lipid-lowering treatment (statins, fibrates or others such as omega3 polyunsaturated fatty acids or ezetimibe); antidiabetic drugs distinguishing insulin, all oral antidiabetic drugs, including tubular sodium glucose co-transporter inhibitors (iSGLT2) or glucagon-like peptide-1 (GLP1-RA) receptor agonists.

### 2.3. Laboratory Data and sample management

At baseline (V1) and at the last visits (V2), fasting venous blood and urine samples were collected. A 20 ml of EDTA (Ethylene diamine tetra acetic) blood sample was obtained from all participants. The samples were centrifuged (4000 rpm; 3 ml for 10 min at 4°C) and stored at -80°C until use. For participants undergoing renal replacement therapy with haemodialysis, fasting samples were taken before the mid-week dialysis treatment procedure. Serum, urine, DNA and whole blood samples were stored in freezers of the Nephropathies Research Group (GREN) of the Institut Hospital del Mar d’Investigacions Mèdiques (IMIM) (31) and the Parc de Salut Mar Biobank (MARBiobanc) (32). All samples for clinical analysis were centralized in a single laboratory, the Catalan Reference Laboratory (LRC). The main variables are summarized in tables 1 and 2.

**Table 1.** General characteristic at baseline visits by grades of chronic kidney disease. Grade 1-2, eGFR>90-60 ml/min/1.73m^2^, grade 3 eGFR; 59-30 ml/min/1.73m^2^, grade 4 eGFR; 29-15 ml/min/1.73m^2^ and grade 5 eGFR<15 ml/min/1.73m^2^. CKD: Chronic kidney disease BMI: Body mass index, HBP: High blood pressure, ACEI/ARB: Angiotensin-converting enzyme (ACE) inhibitors/angiotensin II receptor blockers, iDPP4: Inhibitors of dipeptidyl peptidase 4, iSGLT2: Sodium glucose co-transporter inhibitors, GLP1-RA: Glucagon-like peptide-1 receptor agonists, HbA1c: glycosylated hemoglobin, LDL: low density lipoprotein, HDL: high density lipoprotein.

| CKD grade | 1 - 2 | 3 | 4 | 5 | p |
| --- | --- | --- | --- | --- | --- |
| N | 327 | 204 | 70 | 49 | <0.001 |
| Age (years) | 66.2 (8.3) | 73.5 (9.03) | 75 (8) | 67.9 (8.9) | <0.001 |
| Time of diabetes (years) | 15 (8.2) | 17.9 (8.8) | 19.6 (9.7) | 19.8 (8.4) | <0.001 |
| Gender (Male:Female) % | 61.8/38.2 | 61.6/38.2 | 55.7/44.3 | 61.3/38.8 | 0.812 |
| BMI (Kg/m<sup>2</sup>) | 30.2 (5) | 31 (5.2) | 30.5 (4.8) | 28.8 (4.9) | 0.045 |
| Smokers/former smokers % | 24.8/34.5 | 15.2/39.2 | 4.3/45.7 | 4.1/36.7 | <0.001 |
| HBP % | 69.4 | 98.5 | 95.7 | 100 | <0.001 |
| Antihypertensive treatment<br>ACEI/ARB/ACEi+ARB % | 32.7/39.7/3.1 | 35.3/45.6/2.9 | 11.4/41.4/5.7 | 14.3/49/0 | <0.001 |
| Cardiovascular events history (%) | 31.5 | 46.1 | 60 | 48.9 | <0.001 |
| Ischemic cardiomyopathy | 12.8 | 26.5 | 34.3 | 28.6 | <0.001 |
| Cerebrovascular disease | 9.5 | 11.3 | 37.1 | 10.2 | 0.822 |
| Peripheral vascular disease | 14.7 | 23 | 12.9 | 22.5 | 0.019 |
| Diabetic retinopathy % | 17.1 | 27.9 | 34.3 | 63.3 | <0.001 |
| Lipid-lowering therapy (statin %) | 71.8 | 80.9 | 85.7 | 85.7 | 0.002 |
| Antidiabetic treatment % |  |  |  |  |  |
| Oral agents/Insulin/combined | 59.3/6.1/33.9 | 43.1/28.4/26.9 | 7.3/58.6/20 | 8.1/79.6/6.1 | <0.001 |
| iDPP4/iSGLT2/GLP1-AR | 7.6/0.3/0.9 | 1.9/0.5/0.5 | 1.4/0/0 | 0/0/0 | 0.808 |
| eGFR (mL/min/1.73m<sup>2</sup>) | 82.2 [24.1] | 42.8 [13.2] | 23.5 [19.6] | 9.14 (3.75) | <0.001 |
| Albuminuria/Cr (mg/g) | 9.5 [53.3] | 85.8 [434] | 465 [1574.7] | 1158 [3210.8] | <0.001 |
| HbA1C (%) | 7.7 (1.3) | 7.8 (1.3) | 7.6 (1.2) | 7 (1.1) | 0.004 |
| Cholesterol (mg/dL) |  |  |  |  |  |
| Total | 177.8 (36.5) | 168 (34) | 169 (41.5) | 146.2 (40.3) | <0.001 |
| LDL | 101.3 (30.9) | 92.4 (27.4) | 98 (35.6) | 72.3 (31.7) | <0.001 |
| HDL | 47.7 (12.5) | 46.2 (13.2) | 46.6 (14.3) | 46.5 (15.1) | 0.665 |
| Triglycerides (mg/dL) | 129 [91.7] | 144 [86] | 141 [93] | 125 [60] | 0.072 |
| Uric acid (mg/dL) | 5.5 (1.3) | 6.7 (1.7) | 7.2 (2) | 5.3 (1.7) | <0.001 |
| Hemoglobin (mg/dL) | 13.6 [2.1] | 12.6 [1.88] | 11.5 [1.45] | 12.1 [2] | <0.001 |

**Table 2:** General characteristics at baseline visits and at the end of follow-up visit. BMI: Body mass index, ACEI/ARB: Aangiotensin-converting enzyme (ACE) inhibitors/angiotensin II receptor blockers, iDPP4: Inhibitors of dipeptidyl peptidase 4, iSGLT2: Sodium glucose co-transporter inhibitors, GLP1-RA: Glucagon-like peptide-1 receptor agonists, eGFR: estimated glomerular filtration rate, HbA1c: glycosylated hemoglobin, LDL: low density lipoprotein, HDL: high density lipoprotein.

|  | **First Visit**<br>**2012-2015** | **Last Visit**<br>**2017-2020** |
| --- | --- | --- |
| **N** | 650 | 442 |
| **Age (years)** | 69.6 (9.3) | 73 (8.9) |
| **Sex (male/female.%)** | 61.1/38.9 | 61.5/38.8 |
| **BMI (Kg/m<sup>2</sup>)** | 30.4 (5.08) | 29.75 (5.3) |
| **Time of diabetes (years)** | 16.8 (8.9) | 20.05 (8.5) |
| **Family history of diabetes (%)** | 47.1 | 53.1 |
| **Cardiovascular risk factors history** |  |  |
| Smokers/former smoker (%) | 18/37.4 | 15.8/41.4 |
| High Blood Pressure (%) | 91.4 | 90.9 |
| Dyslipidemia (%) | 77.2 | 73 |
| **Cardiovascular events history (%)** | 40.5 | 41.17 |
| Myocardial infarction (%) | 20.6 | 21.3 |
| Cerebrovascular disease (%) | 10.5 | 14.3 |
| Peripheral vascular disease (%) | 19.8 | 20.4 |
| **Diabetic retinopathy (%)** | 25.8 | 30.5 |
| **Current medication** |  |  |
| **Antihypertensive treatment** |  |  |
| ACEI/ARB/ACEI+ARB (%) | 29.8/40.6/2.8 | 31/37.8/2.3 |
| Others (Calcium-antagonist, β-blocker, diuretics) (%) | 78 | 80.8 |
| **Lipid-lowering therapy (%)** |  |  |
| Statins | 72.6 | 68.1 |
| Fibrates | 10.1 | 6.8 |
| Other | 3.4 | 7.8 |
| **Antidiabetic treatment** |  |  |
| Oral agents only (%) | 46.3 | 41.2 |
| DPP4i/SGLT2i/GLP-1a (%) | 6.1/0.3/0.6 | 21/5.9/4.3 |
| Insulin only (%) | 24.3 | 21 |
| Oral agents + insulin (%) | 28.3 | 34.6 |
| Diet (%) | 1.1 | 1.1 |
| **Serum Creatinine (mg/dl)** | 1.12 [0.81] | 1.11 [0.78] |
| **eGFR (ml/min/1.73mt2)** | 60.4 [46.5] | 54.5 [48.6] |
| **Urinary albumin/creatinine (mg/gr)** | 34.1 [221.5] | 32.6 [212] |
| **Hemoglobin (gr/dl).** | 13.2 (4.9) | 13.4 (5.7) |
| **HbA1c (%)** | 7.6 [1.75] | 7.2 [1.67] |
| **Uric acid (mg/dl)** | 6.25 (1.71) | 6.05 (1.8) |
| **Total Cholesterol (mg/dl)** | 173.3 (37.5) | 166.7 (42.5) |
| **LDL-cholesterol (mg/dl)** | 97.7 (31.4) | 95.4 (32.7) |
| **HDL-cholesterol (mg/dl)** | 47.1 (37.1) | 47.6 (15.1) |
| **Triglycerides (mg/dl)** | 153.9 (79.8) | 162.8 (89.9) |

Renal function was measured as estimated glomerular filtration rate (eGFR) from calibrated serum creatinine using the Chronic Kidney Disease Epidemiology Collaboration (CKD-EPI) equation (33). Moderate albuminuria was defined as a urine albumin-to-creatinine ratio (ACR) of 30-299 mg/g, and severe albuminuria was defined as a urine ACR of 300 mg/g or greater. DKD was defined as: eGFR<60 ml/min/1.73m2 and albuminuria >300 mg/g or albuminuria 30-299 mg/g and DR, regardless the eGFR. Patients were classified based on the degree of kidney disease following the KDIGO guidelines as grade 1-2 if eGFR>90-60 ml/min/1.73m^2^, grade 3 if eGFR; 59-30 ml/min/1.73m^2^, grade 4 if eGFR; 29-15 ml/min/1.73m^2^ and grade 5 if eGFR<15 ml/min/1.73m^2^ (34).

#### Novel molecules and biomarkers

Alongside with the conventional epidemiological phenotypes assessed by questionnaires, clinical visits, analytical and medical reports, the GenoDiabMar registry also benefits from high-throughput techniques to assess new biomarkers related to T2D complications.

#### Metabolomic profiles

Metabolic profiling of 227 metabolic traits, 143 metabolite concentrations, 80 lipid ratios, 3 lipoprotein particle sizes and a semi-quantitative measure of albumin, were determine by Nightingale Health Ltd. (Helsinki, Finland) using a targeted NMR (nuclear magnetic resonance) spectroscopy platform that has been extensively applied for biomarker profiling as previously described (35–39). The metabolomics GenoDiabMar data has been used in a collaborative research study with other three European cohorts TwinsUK (40), KORA (41) and YoungFinns (42), that allowed us to detect metabolomic profiles associated with kidney function, and to identify similarities and discordances between diabetic and non-diabetic populations (26). The registry has metabolomic data from both, the initial and final follow-up visit samples. This may help to assess not only the prognostic value of different metabolic profiles on kidney function and other diabetes complications, but also to rule out the fortuity or causality of the findings.

#### Immunoglobulin G (IgG) glycans analysis

Glycosylation is the most abundant and diverse form of the post-translational modifications of the proteins. Glycans not only play a structural, but also an important functional and regulatory role, and participates in most physiologic process. Immunoglobulin G (IgG) is involved in infectious, autoimmune and inflammatory processes (43–46). Variations in its glycans structures influence the effector function of IgG, modulating its immune response as pro-inflammatory or anti-inflammatory. IgG glycans have been associated with a high variety of conditions, including CKD (27,47). All the GenoDiabMar participants have 76 IgG glycans profiles analysed by high performance liquid chromatography (UPLC) in GENOS, Glycoscience Research laboratory (Zagreb, Croatia). Additionally, to improve our understanding of the role of IgG glycosylation in the evolution of kidney function and antibody-mediated rejection in kidney transplantation, the profiles of IgG glycans have been determined with the same UPLC technique in a sub-population of 248 kidney transplant patients.

#### Other biomarkers available

The cohort also has information on emerging biomarkers of cardiovascular damage, measured in a targeted way, to study their role in kidney damage associated with T2D and how these are influenced by kidney function and albuminuria. In this way, we have carried out the determination of Galectin 3 and Succinate in a sub-set of participants. **Galectin 3** (Gl3) is a β-galactoside–binding lectin that has emerged as a key regulator of inflammation and fibrosis (48–51). Higher Gl3 levels have been linked to an increased CV morbid-mortality risk in the general population. It has been correlated to ischemic cardiomyopathy and lower eGFR in non-diabetic cohorts (52). However, the role of its circulating levels on kidney function in T2D populations has not been fully explored (53,54). In the GenoDiabMar registry, we determined the circulating serum levels of this biomarker using ELISA techniques in a sub-set of 369 patients from the nephrology consultations, which could help to understand its relationship with renal function and its prognostic value in this population. Along the same line, circulating levels of **succinate** were also measured by fluorometric assay (EnzyChromTM Kit) (55) in 602 participants of the cohort. The succinate is a metabolite produced by both the microbiota and the host. It is an intermediate component of the tricarboxylic acid cycle, involved in the formation and elimination of reactive oxygen species that has been linked with hypertension, ischemic heart disease, T2D and obesity (56–59). Thus, in recent years it has been postulated as a marker of high CV risk. Experimental studies suggest that its tissue accumulation and the signalling of its receptor (SUCNR1) activates the cellular response that triggers kidney damage in diabetes and increases the release of renin (60). Nevertheless, little is known about its practical involvement in the clinic and has never been studied in populations with decrease renal function.

#### DNA banking

To facilitate future genetic studies, all the participants in the registry underwent DNA extraction from the whole blood sample obtained at the baseline visit. DNA was extracted in an automated method by Qiasymphony using the Qiasymphony DSP DNA, kit for whole blood, in the MarBiobank facilities. This DNA bank will facilitate future genomic sequencing analysis projects. This cohort is postulated as ideal for conducting studies to confirm and replicate results in the search for genetic markers associated with complications of T2D in larger cohorts. Likewise, it will allow to broaden holistic evaluation analysis, based on system biology studies.

## RESULTS

### General characteristics

The most relevant clinical characteristics and analytical variables of the cohort are displayed in **Table 1 and Table 2**. A total of 650 participants, 61% men and 39% women, aged 69.56 ±9.31 with a median time of diabetes of 15 [11-21] years, underwent the first visit. Of those, 356 (54.7%) had diabetic kidney disease at baseline and distribution per degree of chronic kidney disease was; G1-2 50.3%, G3 31.4%, G4 10.8% and G5 7.5%. Roughly 5 years later (Last in-person visit), 442 participants with their kidney functioning ended the follow up (Table 2). As expected, the presence of DR was significantly more frequent as glomerular filtration rate worsened and was present in 25.8% at baseline and 30.5% in the last visit. The mean body mass index (BMI) was 30.4 (± 5.08) Kg/m^2^. Among the participants, 47.1% had a history of associated family history of T2D. 18% of the patients were active smokers, while 37.4% were ex-smokers. It should be noted that as the degree of CKD worsens, we found a significantly lower percentage of smokers. The prevalence of arterial hypertension was high, with 91.4% of the population being affected and 77.2% had dyslipidaemia. Regarding the history of previous CV events, 20.6% had a history of ischemic heart disease, with a higher prevalence peak in individuals with grade 4 CKD, 10.5% had suffered from cerebrovascular disease and 19.8% had peripheral vascular disease, both ailments again, with a higher prevalence in grade 4 CKD. In respect of anti-hypertensive treatment: 29.8% received angiotensin-converting enzyme inhibitors (ACEI), 40.6% angiotensin receptor blockers (ARB) and 2.8% a combination of ACEI and ARB. As detailed in the tables, as glomerular filtration rates worsen, inhibitors of the renin-angiotensin system usage decreases, with a significant drop in grades 4-5. In addition, 78% of the participants received calcium channel blockers, beta-blockers, or diuretics. 72.6% used statins with a higher prevalence in grade 4 of CKD. Concerning antidiabetic drugs, 46.3% of the participants were taking oral antidiabetic drugs (OADs), 24.3% insulin, and 28.3% combined both treatments (OADs and insulin). As detailed in the tables, the use of OADs decreases significantly as the eGFR worsens, with a clear increase in the insulin usage, being the treatment of choice in 79.6% of patients with grade 5 CKD at baseline visit. Of note, the use of drugs with a demonstrated nephroprotective effect such as SGLT2i or GLP1-RA at the baseline visit (years 2012-2015) was practically anecdotal, with a clear tendency to increase their prescription on the final visit. Albeit, at the beginning of 2020, they were still far from being part of the treatment in most of these patients and, in line as other studies, the actual use of these drugs kept apart from the current clinical practice guidelines (61,62).

Longitudinal data are available for a wide range of clinical and analytical variable as well as for metabolomic and glycomic studies. We assessed the evolution of renal function in the 611 participants with functioning kidneys at the baseline visit (non-dialysis patients). The overall mean annual glomerular filtration loss was -1.2 (−1.8: -0.5 mL/min/1.73m^2^), being different between men and women. Men had a median eGFR higher than women at baseline (66.3 [41.8: 84.3] vs 63.4 [41.7: 86.4] mL/min/1.73m^2^), but the rate of loss of kidney function was significantly lower in women, losing 0.93 (0.40-1.46) less glomerular filtration units per year than men, regardless of albuminuria **Figure 1**. Roughly 17% of the patients experienced rapid progression of renal function, defined as loss of ≥ 5 ml/min/1.73 m^2^/year (34) over the follow-up period, of those 75.2% were men and 24.7% were women.

**Figure 1:**
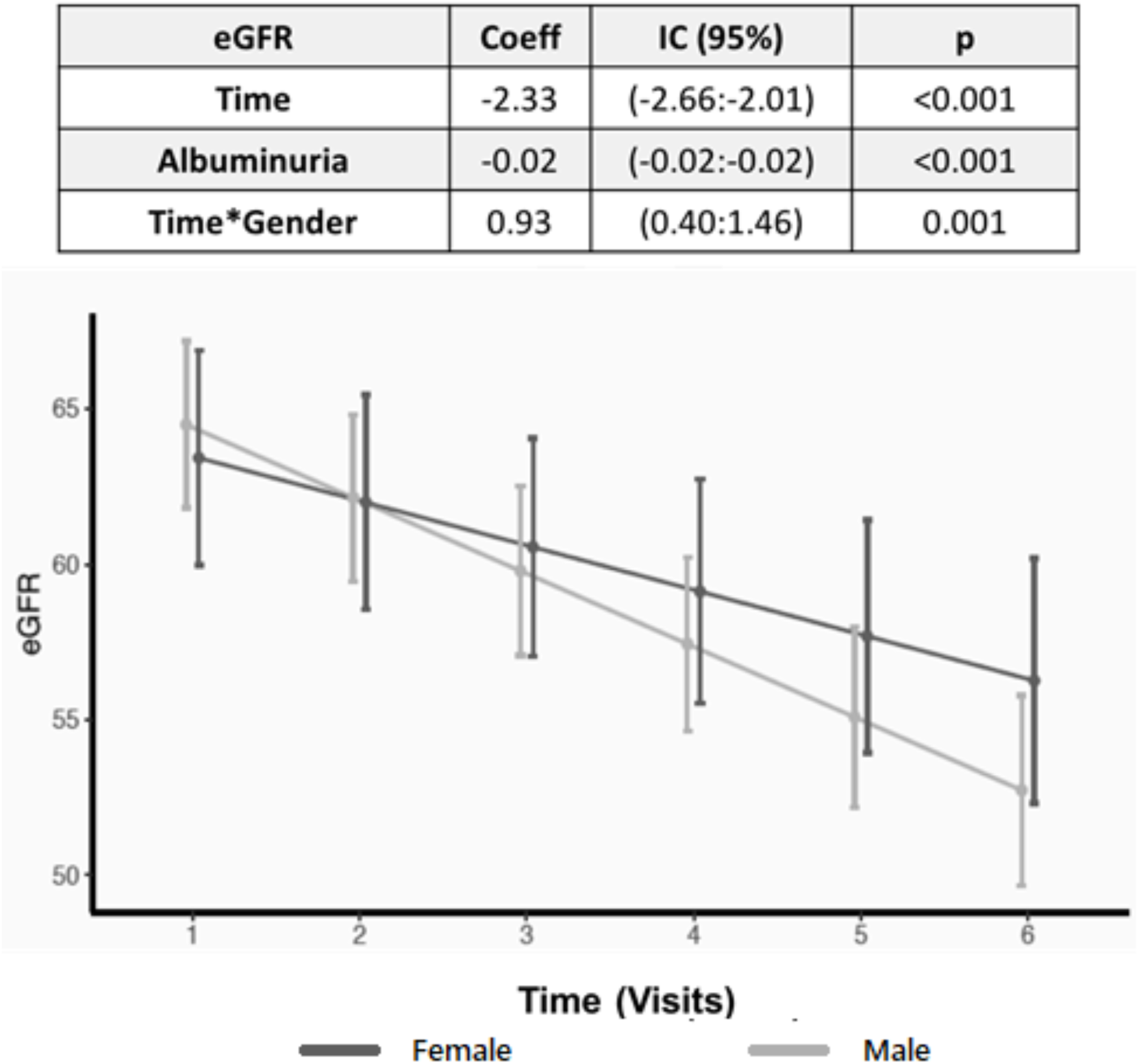
Evolution of estimated glomerular filtration rate (eGFR) per sex, adjusted by albuminuria, in the Mixed Linear Model and its graphic representation of the mean eGFR between sexes.

The parameters influencing a faster progression of renal function were different between sexes. As depicted in **table 3**, the presence of peripheral vascular disease was a risk factor for women but not for men. Also, macroalbuminuria was a significant factor for men but not for women. The AUC for predicting rapid progression by albuminuria was 0.62[0.55:0.68] with a cut-off value of 451 mg/g for men and 0.70[0.62-0.78] with a cut-off value of 18.4 mg/g for women. These findings can help to identify early clinical and analytical risk factors for worse renal evolution in a differential and more personalized way. The characteristics of the evolution of renal function and the differences found between the sexes are like those described in the literature (63–65). This observation reinforces the value of this registry as a population with renal characteristics comparable to other cohorts to be able to carry out collaborative studies. Although this registry was initially started with the objective of studying new biomarkers of kidney injury in T2D, it is a well-characterized population that has detailed information on other micro and macrovascular diabetic complications. During the follow-up period 135 patients (22.1%) endured one or more CV events (**Figure 2**), of which 33.3% fulfilled the established criteria for DKD and 18.4% did not match the DKD criteria at baseline (p<0.001). During the follow-up, 137 patients (22.4%) developed DR or worsening of the previous stablished one (**Figure 2**).

**Table 3:** Risk factors of having rapid progression of kidney function by sex. Multivariate logistic regression model showing the variables of risk of having a rapid decline of renal function, defined as a loss of > 5ml/min/m^2^ of estimated glomerular filtration rate (eGFR) per year, separated by sex.

|  | Men |  | Women |  |
| --- | --- | --- | --- | --- |
|  | *OR (IC95%)* | *p* | *OR (IC95%)* | *p* |
| Age | 1.01 (0.98:1.04) | 0.62 | 1.02 (0.97:1.08) | 0.48 |
| Diabetic retinopathy | 1.18 (0.61:2.23) | 0.61 | 0.99 (0.25:3.33) | 0.98 |
| Time of DM2 | 1.02 (0.99:1.06) | 0.18 | 0.97 (0.91:1.03) | 0.42 |
| BMI | 1.03 (0.96:1.10) | 0.45 | 0.97 (0.89:1.05) | 0.47 |
| Ischemic cardiopathy | 1.02(0.53:1.88) | 0.94 | 1.16(0.31:3.39) | 0.80 |
| Periph. Vascular disease | 0.79 (0.39:1.53) | 0.49 | **3.32 (1.10:9.57)** | **0.02** |
| Stoke | 1.83(0.85:3.74) | 0.12 | 1.82(0.38:6.21) | 0.41 |
| Albuminuria>300mg/g | **2.40 (1.29:4.44)** | **0.005** | 0.99(0.91:3.73) | 0.99 |
| HbA1c | 0.89 (0.71:1.11) | 0.32 | 1.14 (0.80:1.59) | 0.43 |
| Smoker | 1.03 (0.46:2.30) | 0.94 | 1.15 (0.21:4.97) | 0.86 |
| former smoker | 0.72 (0.37:1.46) | 0.35 | 0.29 (0.02:1.62) | 0.25 |

**Figure 2:**
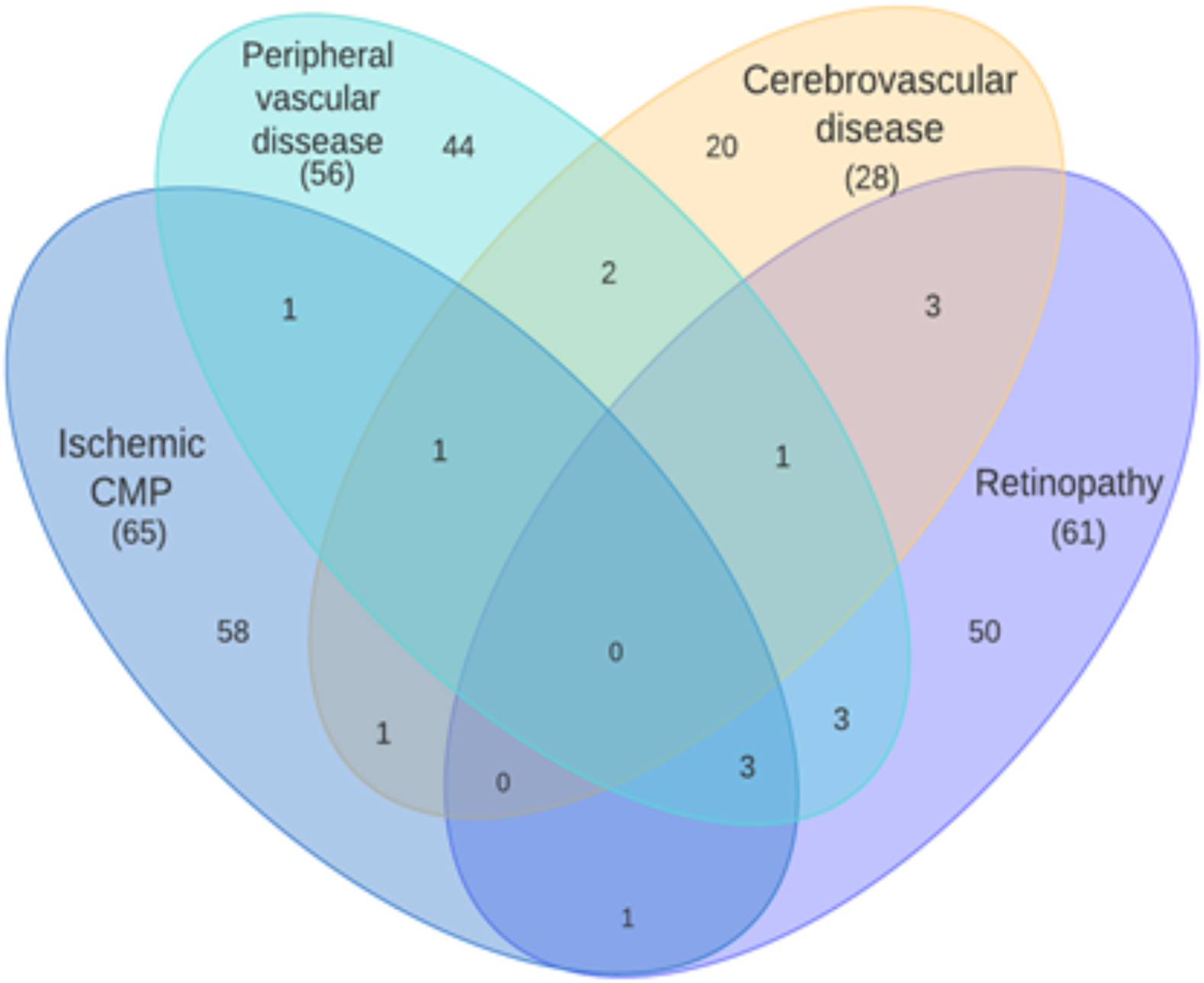
Schematic distribution of the recorded cardiovascular events (Ischemic heart disease, Peripheral vascular disease, Cerebrovascular disease) and new onset of diabetic retinopathy, throughout the follow-up period.

There was an overall mortality of 23%, 38% due to cardiovascular causes and 16% due to cancer. Also, throughout the follow-up, 22 patients (3.6%) started renal replacement therapy and 10 were lost to follow-up (**Figure 3**).

**Figure 3:**
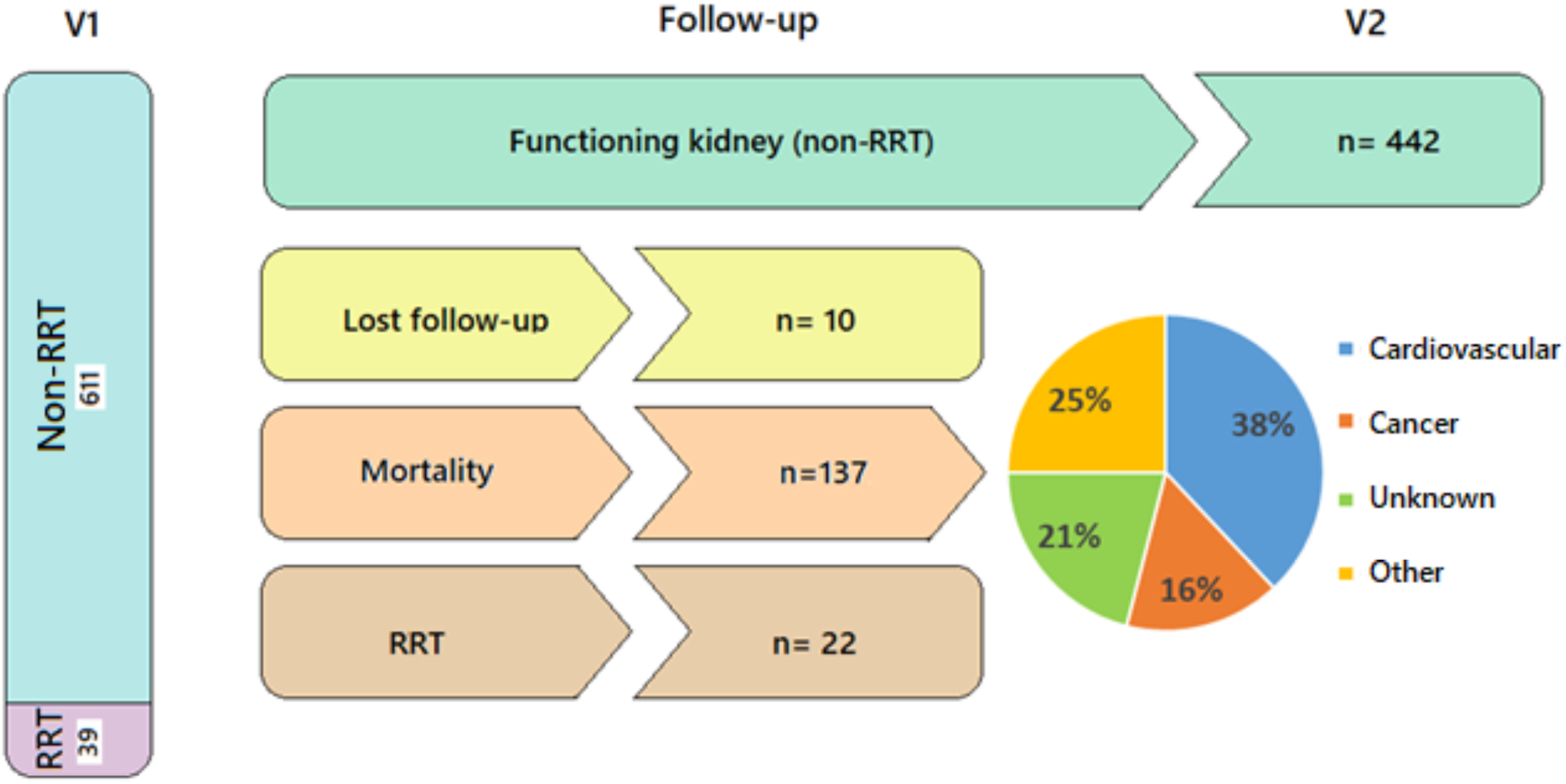
Flow-chart depicts patients’ distribution during follow-up, need of renal replacement therapy (RRT), mortality and it causes.

## DISCUSSION

This is a well characterized registry that gathered longitudinal micro and macrovascular complications of T2D as well as detailed clinical and analytical information. In addition, its main strengths are that includes a collection of baseline and longitudinal follow-up biological samples and covers the entire spectrum of kidney disease, including patients from grade 1 to patients on renal replacement therapy. As they are patients from medical consultations belonging to our healthcare area we have detailed and precise information about them and we can ensure the monitoring capacity. In this way, the participants of this registry are patients from the “*real world”* who sometimes may be excluded from other types of clinical trials or studies, with strict inclusion criteria. One of the limitations of the study is the generalizability of the findings, which is limited to Caucasian subjects. The results of our studies may be validated in multi-ethnic cohorts to evaluate the applicability in broader populations with T2D. We do not have urine samples from all the participants and lack information regarding diet and lifestyle. Also, we do not have kidney histological studies in most of the patients to ensure the renal disease aetiology. Since no other reliable and non-invasive markers have been established, we cannot overcome this limitation but, to minimize misdiagnosis, medical history including ultrasonography and fundoscopy studies were reviewed by two nephrologists.

As described, we have a registry of patients from nephrology and primary care real medical out-patient consults, from which we have registered several clinical and analytical variables for 5 years. In addition, the registry includes serum and urine biobank, DNA bank, as well as data on metabolomics, glycomics and other biomarkers already analysed. Complete list of data and samples available at baseline visit, last visit and annually during follow-up are summarized in **Table 4**.

**Table 4:** Summary available data and samples from different visits and follow-up. V1: First in-person visit, V2: Last in-person visit. HBP: High blood pressure, DL: dyslipidemia. HbA1c: glycosylated hemoglobin, LDL: low density lipoprotein, HDL: high density lipoprotein. NMR: nuclear magnetic resonance spectroscopy, UPLC: high performance liquid chromatography.

|  | **V1. First Visit**<br>**2012-2015** | **Follow-up**<br>**(annually)** | **V2. Last Visit**<br>**2017-2020** |
| --- | --- | --- | --- |
| **Questionnaires** |  |  |  |
| Demographics | ✓ |  |  |
| Time of diabetes | ✓ |  | ✓ |
| Family history of diabetes | ✓ |  |  |
| Cardiovascular risk factors<br>(Smoke, HBP, DL) | ✓ | ✓ | ✓ |
| Cardiovascular events history | ✓ | ✓ | ✓ |
| Current medication<br>(anti-hypertensive, lipid-lowering,<br>anti-diabetics treatments) | ✓ | ✓ | ✓ |
| Renal replacement therapy |  | ✓ | ✓ |
| Mortality |  | ✓ | ✓ |
| **Clinical assessments** |  |  |  |
| Anthropometrics (BMI) | ✓ |  | ✓ |
| Blood pressure | ✓ |  | ✓ |
| Fundoscopy | ✓ |  | ✓ |
| Abdominal ultrasound | ✓ |  |  |
| **Laboratory analysis** |  |  |  |
| Biochemical profile (creatinine,<br>urea, uric acid) | ✓ | ✓ | ✓ |
| Hemogram | ✓ |  | ✓ |
| Lipid profile (total cholesterol, LDL,<br>HDL-cholesterol, triglycerides) | ✓ |  | ✓ |
| Albuminuria | ✓ | ✓ | ✓ |
| HbA1c | ✓ | ✓ | ✓ |
| **Special test** |  |  |  |
| Metabolomic profiles (NMR) | ✓ |  | ✓ |
| IgG glycans profiles (UPLC) | ✓ |  | ✓ |
| Serum Galectin 3 (ELISA) | ✓ |  | ✓ |
| Serum Succinate (fluorometric) | ✓ |  | ✓ |
| **Available samples** |  |  |  |
| DNA bank and whole blood sample | ✓ |  | ✓ |
| Serum samples | ✓ |  | ✓ |
| Urine samples | ✓ |  | ✓ |

We consider that this cohort to be postulated as a great tool for scientific collaboration for studies, whether they are focused on T2D, or whether they are interested in comparing differential markers between diabetic and non-diabetic populations. Furthermore, as we have shown in other collaborative projects, the GenoDiabMar registry can meet the criteria to replicate or meta-analyse results obtained in other cohorts. It should be noted that this registry is part of The Consortium of Metabolomics Studies (COMETS) whose main objective is to create a collaborative network to identify metabolomic markers associated with different phenotypes and pathologies (66). In this way, collaboration with the GenoDiabMar project could open collaborations with other studies worldwide that are part of COMETs.

The main objective of this descriptive publication of our GenoDiabMar registry is to engage researchers in collaborative efforts to advance knowledge of the aetiology, diagnosis, treatment, and prognosis of T2D complications. In that spirit, we invite researchers including those without data of their own, to join us with scientific collaboration proposals.

## Data Availability

All data produced in the present study are available upon reasonable request to the authors

## Acknowledgements

Clara Barrios is funded by grants FIS-FEDER-ISCIII PI16/00620 (Ext 2021) and Strategic Plan for Research and Innovation in Health, CatSalut, PERIS STL008 (2019-2021) to develop clinical and epidemiological studies mainly focused in diabetes and its associations with new biomarkers.

## Author Contributions

Conceptualization, C.B.; methodology, A.S., S.O., A.F., M.V., J.F-N., X.M., validation, C.B. and J.P.; formal analysis, A.S., S.O., X.D., A.C-G and C.B.; investigation, A.S., E.R, E.M and C.B.; resources and funding acquisition, J.P, M.C and C.B; data curation, A.S., S.O., X.D., A.C-G and C.B.; writing—original draft preparation, A.S, S.O. and C.B.; writing—review and editing, all authors; supervision, C.B.; All authors have read and agreed to the published version of the manuscript.”

## Conflicts of Interest

The authors declare no conflict of interest. The funders had no role in the design of the study; in the collection, analyses, or interpretation of data; in the writing of the manuscript, or in the decision to publish the results.

## Notes

### Competing Interest Statement

The authors have declared no competing interest.

### Author Declarations

Ethics committee/IRB of Institut Hospital del Mar d'Investigacions Mediques gave ethical approval for this work

